# Using remotely delivered Spring Forest Qigong™ to reduce neuropathic pain in adults with spinal cord injury: Protocol of a quasi-experimental clinical trial

**DOI:** 10.1101/2022.03.09.22271844

**Authors:** Ann Van de Winckel, Sydney Carpentier, Wei Deng, Lin Zhang, Ricardo Battaglino, Leslie Morse

## Abstract

**Background:** About 69% of Americans living with spinal cord injury (SCI) suffer from long-term debilitating neuropathic pain, interfering with the quality of daily life. Neuropathic pain is refractory to many available treatments –some carrying a risk for opioid addiction– highlighting an urgent need for new treatments. In this study, we will test our hypothesis that Spring Forest Qigong^TM^ will reduce SCI-related neuropathic pain by improving body awareness. We will determine whether remotely delivered Qigong is ***feasible*** and we will collect data on neuropathic pain, and other reported associations with pain such as spasms frequency and/or severity, functional performance, mood, or body awareness.

**Methods:** In this quasi-experimental pilot clinical trial study, adults with SCI will practice Qigong at home with a 45min video, at least 3x/week for 12 weeks. The Qigong practice includes movements with guided breathing and is individualized based on functional abilities, i.e., the participants follow along with the Qigong movements to the level of their ability, with guided breathing, and perform kinesthetic imagery by focusing on the feeling in the whole body as if doing the whole-body Qigong movement while standing. The highest, average, and lowest neuropathic pain ratings perceived in the prior week will be recorded weekly until the 6-week follow-up. The other outcomes will be collected at 5 time points: at baseline, midway during the Qigong intervention (6 weeks), after the Qigong intervention (12 weeks), after a 6-week and 1-year follow-up. Rate parameters for the feasibility markers will be estimated based on the participants who achieved each benchmark.

**Discussion:** The University of Minnesota (UMN)’s Institutional Review Board (IRB) approved the study (IRB #STUDY00011997). All participants will sign electronic informed consent on the secure UMN REDCap platform. The results will be presented at academic conferences and published in peer-reviewed publications.

**Trial registration:** ClinicalTrial.gov registration number: NCT04917107, first registered 6/8/2021, https://clinicaltrials.gov/ct2/show/NCT04917107.

## INTRODUCTION

### Background and rationale {6a}

About 69% of the 299,000 Americans living with spinal cord injury (SCI) suffer from debilitating neuropathic pain, interfering with general activity, mobility, mood, sleep, and quality of life.[1–3] Yet, neuropathic pain is refractory to many treatments, and some medications carry a risk for opioid addiction, highlighting a need for new treatments.[4]

SCI dramatically alters the inflow of sensory information.[5–7] The brain uses a prediction framework in which predictions of sensory input are compared to actual sensory input.[5–7] This disconnect between predicted and actually perceived sensory information leads to altered brain circuitry related to sensory information, including increased pain sensation, which in turn, drives the neuropathic pain perception.[5–7]

Concurrently, recent studies showed that SCI-related brain alterations impact body awareness in addition to pain perception.[8–11] Mind and body approaches –known for training body awareness– have been shown to reduce pain in populations with chronic low back pain and fibromyalgia, but evidence of its effectiveness in adults with SCI is limited.[12–20] Adaptive Yoga vs. waitlist group decreased depression and improved self-compassion, but not pain in adults with SCI.[19] A seated Tai Chi program (single-arm study) was well-tolerated in adults with SCI, reduced pain, and improved emotional and physical well-being immediately after each session.[20] However, the weekly in-person classes were associated with a 60% drop-out, and only included adults with paraplegia were included. While these are promising results, more studies are needed.

Qigong –a mind and body approach that incorporates gentle body movements, paired with a focus on breathing and body awareness to promote health and wellness– is accessible for adults with SCI due to the simple, gentle movements, which can be done in a sitting or lying position or by doing kinesthetic imagery of the movements in addition to active movements to the level of their ability.[15, 21] Practicing Qigong has been recognized to improve posture, body awareness, and emotional balance.[15] Some Qigong programs are available online, which helps make the practice accessible for adults with SCI with transportation issues or who live too far away from Qigong centers.

### Objectives {7}

In this quasi-experimental study, we will test our hypothesis that the remotely delivered Qigong will reduce SCI-related neuropathic pain by improving body awareness. If our hypothesis is validated, the outcome of our work will be the first step in demonstrating a potentially effective therapy for adults with SCI-related neuropathic pain. To test this hypothesis, we will determine whether 12 weeks of remote Qigong is ***feasible and well-tolerated*** and whether it reduces neuropathic pain in adults with SCI. Given the reported associations with pain, we will assess whether Qigong practice reduces spasms frequency and/or severity, and improves function, mood, and body awareness. The feasibility of the 6-week follow-up will investigate how long potential Qigong effects remain without practicing. After the 6 weeks, participants can resume practice at will. The feasibility of the 1-year follow-up will examine possible behavioral changes of participants in practicing Qigong in their daily life.

### Trial design {8}

The study flow chart of this quasi-experimental pre-post design is presented in ***Figure 1***. Participants with SCI-related neuropathic pain and with complete or incomplete paraplegia or tetraplegia will perform 12 weeks of “Five Element Qigong Healing Movements” of Spring Forest Qigong^TM^ with an online video at home, at least 3x/week, 41 min/session. *Individual weekly check-ins* (15-30min) are organized during the Qigong practice to address questions and demonstrate movements over Zoom if needed. Qualitative information will be collected about their adherence, adverse outcomes if any, perceptions, effects, and satisfaction with the Qigong practice. During the whole study period, we will record weekly the highest, average, and lowest neuropathic pain intensity ratings perceived in the prior week, the dose of neuropathic pain medication, recent illnesses, health care utilization, and/or recent hospitalizations, and any activity that would indicate community integration.

**Figure 1.**
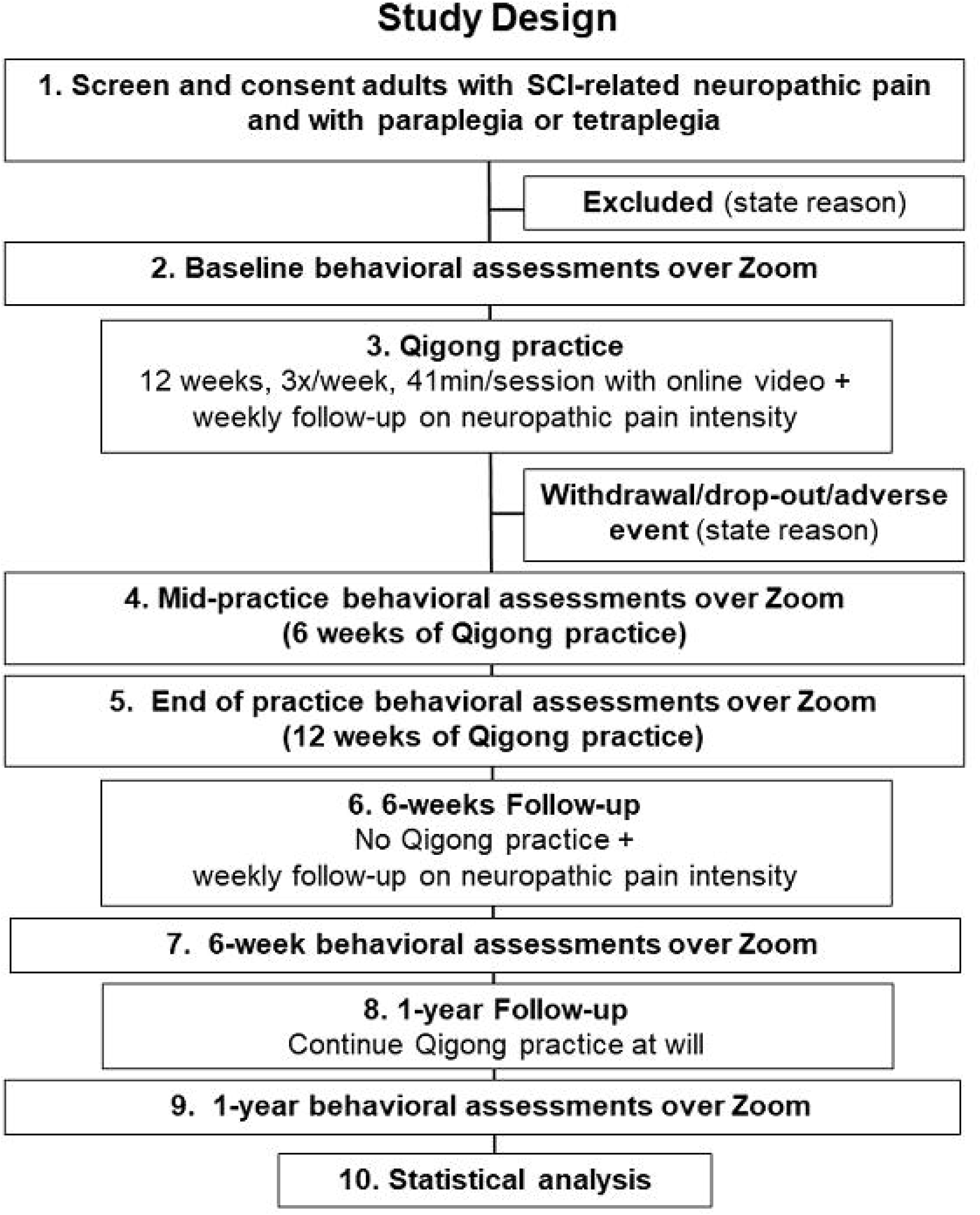
Study flow chart

The outcomes associated with pain will be collected at 5 time points: at baseline, midway during the Qigong intervention (6 weeks), after the Qigong intervention (12 weeks), and after the 6-week and 1-year follow-up. During the 6-week follow-up, participants will not practice Qigong but receive regular standard of care as needed. After the 6-weeks, the participants can restart their Qigong practice at the frequency of their choice so we can evaluate any change in behavior occurring when no instructions are given. Participants will receive $100 upon study completion, through a Greenphire ClinCard. The Qigong introduction class and video access will be paid for by the study.

## METHODS: Participants, intervention and outcomes

### Study setting {9}

Since this is a remote-only study, participants practice at home or in any location of their choice with a Wi-Fi connection.

### Eligibility criteria {10}

We plan on recruiting 18 adults with SCI-related neuropathic pain, who are 18–75 years old, with SCI of ≥ 1 year, medically stable with complete or incomplete paraplegia (T1 and below) or tetraplegia (C4 and below), and with the highest level of SCI-related neuropathic pain ≥3 on the Numeric Pain Rating Scale.[22]

We estimate the average age of participants to be 43 years old.[23], and the sex ratio to be 4:1 (men:women).[4, 23] About 24% of SCI occur among non-Hispanic Blacks.[4, 23] Of note, Minnesota has a 79.10% non-Hispanic white population and 7.00% non-Hispanic Blacks.[24] The exclusion criteria are uncontrolled seizure disorder; cognitive or communicative impairments that prevent participants from following directions or learning; ventilator dependency; other major medical complications; treatments for neuropathic pain other than medication.

### Patient and public involvement

Adults with SCI-related neuropathic pain living in the community were invited to share their perception of ‘priority research’ (in this case, treatments for neuropathic pain that are not pharmaceutical or invasive), feasibility and relevant outcomes (listed in this study), assessment, and study duration. They preferred remote interventions because many adults with SCI have transportation issues and some live rurally or out of state.

### Who will take informed consent? {26a}

Eligible participants will have a virtual visit through the UMN secure Zoom conference ability with the study staff to review and address questions about the combined HIPAA/informed eConsent.

### Additional consent provisions for collection and use of participant data and biological specimens {26b}

The eConsent will be signed through a UMN Research Electronic Data Capture (REDCap) link. REDCap uses a MySQL database via a secure web interface with data checks to ensure data quality during data entry. All IRB rules and HIPAA rules protecting the participants’ rights and confidentiality of records will be followed.

## INTERVENTIONS

### Explanation for the choice of comparators {6b}

Because this is the first study on Qigong in adults with SCI-related neuropathic pain, we decided on a quasi-experimental design and did not include a control group or a sham version of Qigong.

### Intervention description {11a} : Qigong intervention

A Qigong Master from the Spring Forest Qigong^TM^ Center (Minnesota) will teach the introductory class of the “Five Element Qigong Healing Movements” (6h) over Zoom. These movements can be done lying or sitting, and thus, they are suitable for adults with SCI with paraplegia or tetraplegia.[15, 21]

Additionally, Dr. Van de Winckel developed specific kinesthetic imagery instructions to allow participants with all levels of mobility to participate maximally in the Qigong practice. The participants will be instructed to actively move however much they comfortably can and perform kinesthetic imagery. The instructions of the kinesthetic imagery are to focus on imagining the feeling of performing the entire whole-body movement standing up while focusing on breathing and body awareness and to imagine the ‘feeling’ of the contact of the floor with the soles of their feet and of the flow of the movements, rather than to ‘visualize’ the whole-body movement. If reproducing this feeling is difficult, participants will be asked to associate positive memories from before their SCI with this imagined standing posture, e.g., positive memories of feeling the warm sand under the soles of their feet on the beach or the feeling of grass under the soles of bare feet in the garden.

After the introductory class, participants will receive a password-protected website link to practice for 12 weeks.

#### To address the feasibility outcome

the Qigong website automatically monitors the days and duration that videos are accessed, which allows us to objectively track adherence. The Spring Forest Qigong^TM^ center will provide the weekly logs to the principal investigator. Since participants will log in with their ID code given at the start of the study, no personal information is transmitted to the website or the center.

#### Qigong Practice

Qigong Master Chunyi Lin, MS in holistic healing, developed the video “Five Element Qigong Healing Movements”, and guides the participant on the online video through each of the five gentle horizontal and vertical arm and leg movements performed with guided breathing.[13, 25] As defined by Traditional Chinese Medicine, each of the five movements connects to a specific natural element (Fire, Earth, Metal, Water, and Wood). Each element is associated with two meridians, with a specific organ system, and with specific emotions, which have a positive and negative counterpart. When practicing the Five Element Qigong Healing Movements, the focus is directed only on positive emotions. The explanation of the five movements (***Figure 2***) is detailed below.[25]

**Figure 2.**
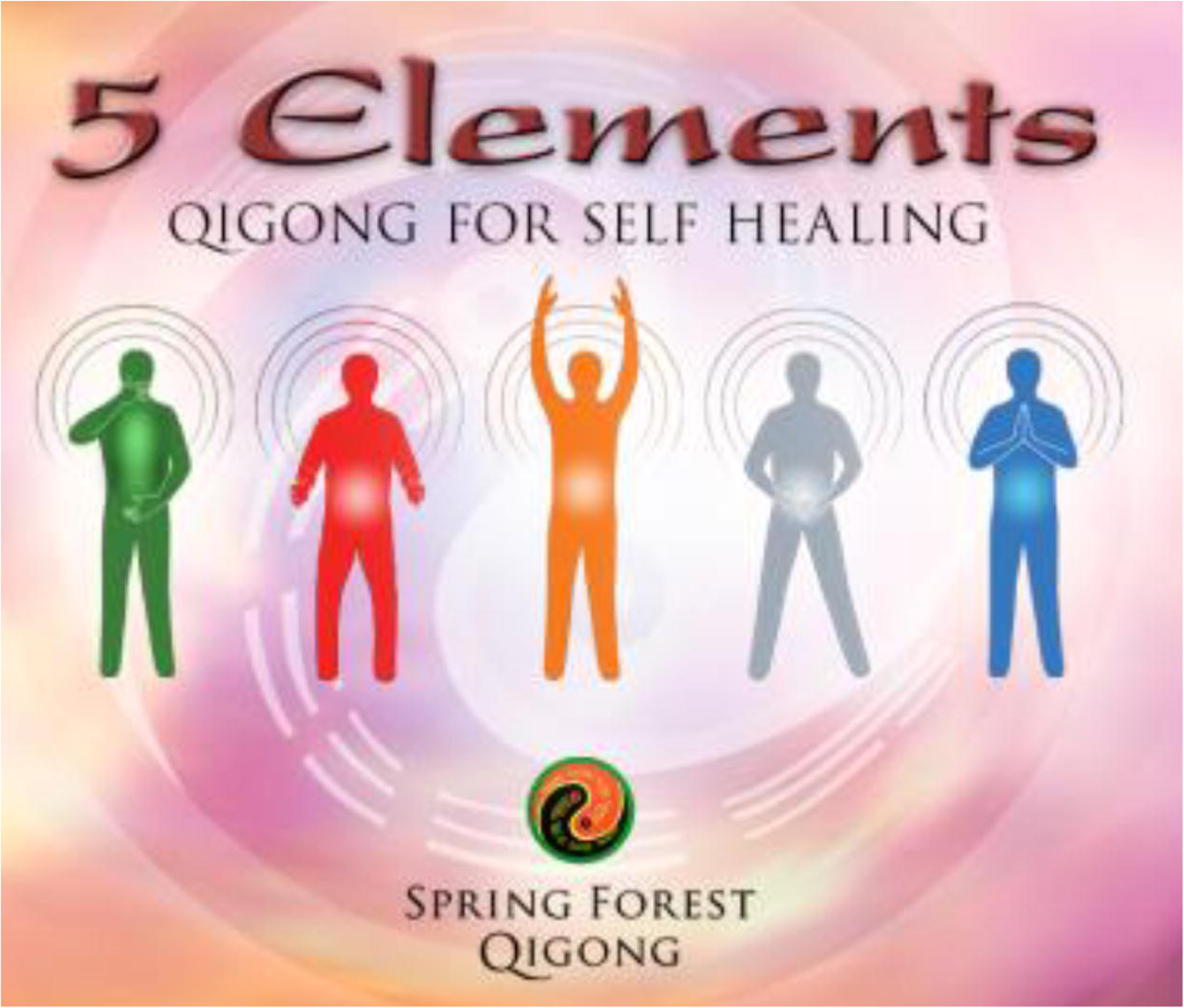
Depiction of the Five Element Qigong Healing Movements from Spring Forest Qigong^TM^. The five movements are presented from left to right. Movement 1: Moving of Yin and Yang; Movement 2: Breathing of the Universe; Movement 3: Connecting with Heaven & Earth; Movement 4: Connecting with Your Body’s Energy; Movement 5: Connecting with Your Heart’s Energy. Details of how the different movements are performed are presented in the text.

##### Preparation phase

The practice starts with smiling and focusing on the heart. The participants bounce for one minute with their arms relaxed next to the body, and another minute with their arms in the air. Master Lin then guides the participants through various cupping and tapping movements to open the energy channels related to each of the five organ systems. The preparation phase ends with setting an intention for healing. Then the five movements are performed for 5 minutes each.

##### Movement 1: Moving of Yin and Yang

This movement helps the liver system and is connected to the Wood element. The liver system is associated with the emotion of happiness (or its negative counterpart: anger). Movement 1 is performed by using the elbows to have the arms make an elliptical movement. The movement starts by moving the right hand out and down to the bottom of the torso, while the left hand moves in (closer to the body) and up to the face. While doing this movement, the focus is on the emotion of happiness and body sensations. After five minutes, the hands are brought down in front of the Lower Dantian, which is an energy center about the width of 3 thumbs below the navel and 3 thumbs inward. The participants focus on the Lower Dantian and take a deep breath three times.

##### Movement 2: Breathing of the Universe

This movement helps the heart system and is connected to the Fire element. The heart system is associated with the emotion of joy (or its negative counterpart: hatred). After Movement 1, on the fourth breath, the participants inhale, bend their knees and open their hands to the sides. With the exhale, the hands come closer again and the participants straighten their body again. Participants focus on the emotion of joy and body sensations.

##### Movement 3: Connecting with Heaven & Earth

This movement helps the stomach and spleen systems and is connected to the Earth element. The stomach and spleen systems are associated with the emotion of peace and groundedness (or its negative counterpart: anxiety and thinking). Movement 3 activates the triple heater, which is an energy system that helps the internal organs, glands, and the lymph system. After Movement 2, the participants lower their hands to their sides and then move their hands up from both sides in the same way as they would hold a ball above their head. The hands are kept above the head during the entire duration of this movement. The participants inhale and bend their knees to lower the body slowly. As they exhale, they straighten their body. Participants focus on the emotion of peace and groundedness and body sensations.

##### Movement 4: Connecting with Your Body’s Energy

This movement helps the lung system and is connected to the Metal element. The lung system is associated with the emotion of contentment (and its negative counterpart: depression and sadness). Movement 4 directs energy to a point called Hui Yin at the center of the pelvic floor, 0.5 inch in front of the anus. This is where the female energy channels gather together. Performing this movement is beneficial for the reproductive organs in women and prostate function in men. The participants bring both hands down and connect the tips of their fingers forming a heart shape in front of their navel. The participants keep a light pressure on the tips of their fingers to stimulate the heart’s energy. The participants bend the knees while keeping the spine and shoulders relaxed and straight and then spread the legs slightly by moving the right foot half a step out. While participants inhale, they shift the body weight to the left, putting 70% of the weight on the left leg and holding this position for three seconds. As they exhale, they switch the body weight and put 70% of the weight on the right leg. They hold this position for three seconds. The participants keep shifting their body weight in this way from side to side. During this movement, participants focus on the emotion of contentment and their body sensations.

##### Movement 5: Connecting with Your Heart’s Energy

This movement helps the kidney system and is connected to the Water element. The kidney system is associated with the emotion of gratitude and thankfulness (and its negative counterpart: fear). Movement 5 helps to move energy to the Bai Hui point on the top of the head. The participants adjust their feet so they are standing with the feet shoulder-width apart. They put the hands together palm to palm in front of the chest while keeping a light pressure on the fingertips to stimulate the heart’s energy. During the inhalation, the participants slowly bend forward at the waist. This is a small movement. With the exhale, the participants straighten back up. While doing this movement the participants focus on the emotion of gratitude and body sensations.

##### The Ending: Harvesting of Qi

The participants rub the hands together, palm to palm, and then massage the face and ears, comb through the hair, followed by cupping and tapping parts of the body. They then make gentle dolphin movements with the neck and body, and after more cupping, the practice ends by lifting and dropping the heels nine times.

### Criteria for discontinuing or modifying allocated interventions {11b}

The risks of Qigong practice are considered to be minimal and expected to be limited to mild transient discomfort. Stopping rules and subsequent removal from the study will be applied when a participant is found to be intolerant to the required study procedure at any time point; serious adverse events at any time point; develops an intercurrent illness that would, in the judgment of the principal investigator, affect the assessment of clinical status to a significant degree; or requests to withdraw from the study. Serious adverse events will be reported as per IRB and Good Clinical Practice requirements. In sum, because of the low-intensity Qigong movements or kinesthetic imagery, we anticipate that this trial will be safe and well-tolerated. Thus, we do not anticipate terminating the trial due to safety or tolerability issues.

### Strategies to improve adherence to interventions {11c}

We will organize weekly phone calls to address questions about the Qigong practice and demonstrate movements over Zoom if needed. Qualitative information will be collected about their adherence, adverse outcomes if any, perceptions, effects, and satisfaction with the Qigong practice.

### Relevant concomitant care permitted or prohibited during the trial {11d}

Concurrent practice of other mind-body approaches is not permitted. Their regular standard-of-care (any medication intake) as well as health care appointments will be maintained to ensure adequate care for other SCI-related symptoms, but appointments for neuropathic pain (e.g., osteopathy) will not be scheduled or permitted during the study.

### Provisions for post-trial care {30}

All assessments and practice are delivered remotely. The study does not provide compensation for research-related injuries or unexpected urgent care.

### Outcomes (12}

#### Feasibility benchmarks

Our feasibility markers are consistent with models and guidelines for intervention development.[26, 27] Based on prior publications,[28–31] we will assess the following *a-priori* feasibility markers.

| Feasibility item & details | Benchmark |
| --- | --- |
| ***Recruitment/enrollment***<br>Assess recruitment rates (screens and enrollment, sex/racial outcomes).<br>Evaluate the recruitment rate for complete and incomplete SCI. | The proportion of potential participants contacted who agreed to participate: 80% is excellent and 70% good.<br>Ratio of complete/incomplete SCI. We estimate recruitment of 30-40% of adults with complete, 60-70% with incomplete SCI.<br>Reasons for (not) participating will help delineate facilitators and barriers to participation. |
| ***Feasibility and acceptability of the study design***<br>Drop-out rates | Proportion of people withdrawing from the study and the reason for withdrawing. A proportion of 90% staying in the study is excellent, 80% good (i.e., 20% drop-out). |
| ***Feasibility, adherence, and acceptability of the Qigong practice***<br>Assess intervention adherence, average Qigong practice; at least 3x/week (123min). | Weekly/ monthly check-ins (reporting min/week of practice) to monitor adherence. The website also logs the day, time, and duration of video access.<br>Feasibility: Excellent: >80% participants practice at least 2x/week; good ≥70%.<br>Drop-out due to lack of interest will be counted as general attrition: 10% drop-out is excellent, 20% is good. |
| ***Feasibility of adhering to the collection*** | Feasibility is 'excellent' when none of the |
| **of quantitative measures**<br>○ % Adherence in completing all assessments during the study (until 6 weeks follow-up).<br>○ % adherence for the 1-year post-intervention assessments. | questionnaires are fully missing in > 25% of the participants. |
| **Program satisfaction**<br>○ interview about satisfaction with the Qigong practice, and the study. | ○ Excellent satisfaction: ≥75% of participants report being satisfied with the study and Qigong practice; and good if ≥70% of participants are satisfied. We will ask them about satisfaction at the end of the 12 weeks of practice and at the end of the 6-week, and 1-year follow-up. |
| **Program safety**<br>○ Assess adverse events (IRB + Good Clinical Practice reporting guidelines). | ○ Excellent safety: no study-related adverse events; good if there are minimal and mild adverse events (e.g., muscle soreness) linked to Qigong practice in max 10% of participants. We will ask about adverse events on our weekly and monthly check-in calls. |

#### We will collect patient-centered outcome measures on neuropathic pain intensity and symptoms associated with pain

##### Neuropathic pain intensity

The highest, average, and lowest neuropathic pain intensity ratings perceived in the prior week will be recorded using the 0-10 Numerical Pain Rating Scale with 0 being no pain and 10 being the worst pain ever.[22] This scale has been recommended by the mission of the Initiative on Methods, Measurement, and Pain Assessment in Clinical Trials consensus group for use in pain clinical trials and by the 2006 National Institute on Disability and Rehabilitation Research SCI Pain outcome measures consensus group.[32, 33]

##### Location and impact of neuropathic pain; spasms frequency and severity

Other pain data will be collected using the National Institute of Neurological Disorders-Common Data Elements International SCI Pain Basic Data Set Version 2.0.[34] This Data Set also assesses dimensions of pain (i.e., the intensity of pain, location, etc.), and how pain interferes with mood, activity, and sleep. The Brief Pain Inventory elicits self-report of pain intensity and its impact on functioning in the past 24h.[33] Spasms frequency and spasm severity will be evaluated with the Penn Spasm Frequency Scale.[35]

##### Mood

The Patient Health Questionnaire-9 assesses symptoms of depression.[36–40] The State-Trait Anxiety Inventory assesses current symptoms of anxiety (State anxiety) and a generalized propensity to be anxious (Trait anxiety).[41]

##### Quality of life

The World Health Organization Quality of Life Instruments (abbreviated version) assesses physical and psychological health, social relationships, and environment.[42–44] The Satisfaction with Life Scale assesses a person’s happiness with the current quality of life.[45] The Moorong Self-Efficacy Scale assesses self-efficacy related to everyday life activities in adults with SCI.[46–48] The Pittsburgh Sleep Quality Index assesses sleep quality.[49]

##### Body awareness

The Revised Body Awareness Rating Questionnaire assesses how pain or tension in the body is affecting participants’ awareness and function in daily life (e.g., being comfortable when lying down).[50] The Postural Awareness Scale measures awareness of body posture in patients with chronic pain.[51] The Functional Appreciation Scale measures the level of appreciation one has for the functionality of the body.[52] The Multidimensional Assessment of Interoceptive Awareness measures interoceptive body awareness.[53] The Body perception questionnaire-short form measures interoceptive awareness and autonomic reactivity.[54, 55] Participants self-report whether their cardiopulmonary system, bladder, bowel, and sexual function are normal or abnormal with the Autonomic Standards Assessment Form.[56]

##### Function

The Spinal Cord Injury Functional Index[57] measures functional performance across 5 domains: basic mobility, self-care, fine motor function, wheelchair mobility, and/or ambulation.[58] For the Patient Specific Functional Scale,[59] the participants will self-identify goals related to activities in daily life that are important to them. The participants will rate them at each time point between 0 (i.e., unable to do the activity without pain) and 10 (able to do the activity without pain).[59] For the Physical activity 3-day recall questionnaire, the participants describe all activities performed during the past 3 days.[60, 61]

##### Fear of injury due to movement

The Tampa Scale for Kinesiophobia assesses fear of injury due to physical movement.[62]

##### Community integration

The Craig Handicap Assessment and Reporting Technique– Short Form is used to assess community integration.[63]

### Participant timeline {13}

***Table 1*** displays the scheduling of assessments and timeline of each study section.

**Table 1.**
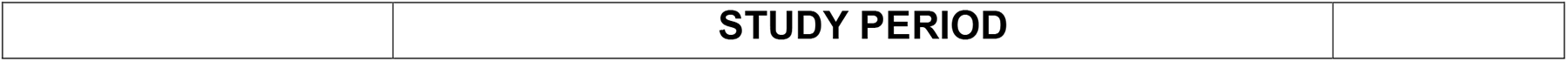
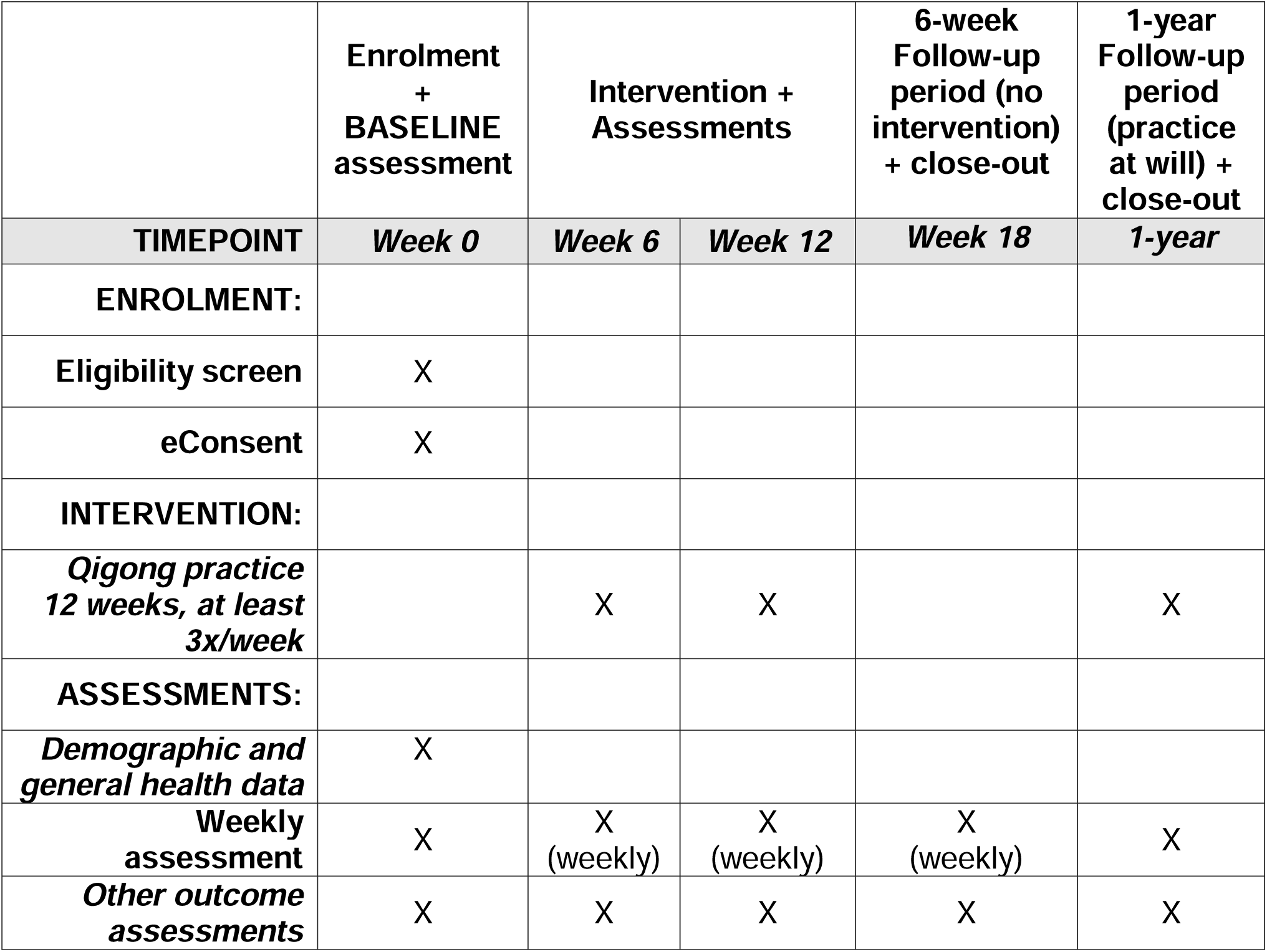
Scheduling of enrolment, intervention, and assessments

|  | STUDY PERIOD |
| --- | --- |

|  | Enrolment<br>+<br>BASELINE<br>assessment | Intervention +<br>Assessments |  | 6-week<br>Follow-up<br>period (no<br>intervention)<br>+ close-out | 1-year<br>Follow-up<br>period<br>(practice<br>at will) +<br>close-out |
| --- | --- | --- | --- | --- | --- |
| TIMEPOINT | Week 0 | Week 6 | Week 12 | Week 18 | 1-year |
| ENROLMENT: |  |  |  |  |  |
| Eligibility screen | X |  |  |  |  |
| eConsent | X |  |  |  |  |
| INTERVENTION: |  |  |  |  |  |
| *Qigong practice*<br>*12 weeks, at least*<br>*3x/week* |  | X | X |  | X |
| ASSESSMENTS: |  |  |  |  |  |
| *Demographic and*<br>*general health data* | X |  |  |  |  |
| Weekly<br>assessment | X | X<br>(weekly) | X<br>(weekly) | X<br>(weekly) | X |
| *Other outcome*<br>*assessments* | X | X | X | X | X |

### Sample size {14}

To our knowledge, this will be the first study evaluating the feasibility of Qigong on neuropathic pain perception. This study will also provide preliminary estimates of effect size, crucial to generate hypotheses for future larger randomized controlled trials. We conducted a preliminary power calculation for the given sample size based on estimates from another body awareness therapy in adults with SCI (currently unpublished). In this prior study, we saw a reduction in neuropathic pain intensity of at least 2.31±2.07 points for the highest, average, and lowest neuropathic pain in the prior week. Assuming the same standard deviation estimate, a total sample size of 18 participants will have over 98% power to detect the same pain reduction of 2.31 points on the numeric pain rating scale (standardized Cohen’s d = 1.12) with a two-sided significance level of 0.05 using a paired t-test, and 80% power to detect a pain reduction of 1.46 points (Cohen’s d=0.72) in this study. With an attrition rate of 30%, the planned sample size will still have 80% power to detect a pain reduction of 1.85 points (Cohen’s d=0.89).

### Recruitment {15}

Participants will be recruited in outpatient locations within the newly funded Spinal Cord Injury Model System Center (Minnesota Regional Spinal Cord Injury Model System) and the community. Specifically, we have collaborators within the Minneapolis VA Health Care System including the Minnesota Paralyzed Veterans; M Health Fairview (Minneapolis); MN SCI associations; Regions Hospital; Courage Kenny Rehabilitation Institute; Get Up Stand Up To Cure Paralysis; Unite2fight paralysis, and Fit4Recovery.

Since this is a remote-only study, we can recruit nationwide. We will use volunteer sampling through fliers, and post study-relevant information on social media and relevant websites. We will also use convenience sampling by having the health care providers in hospitals and organizations provide enrollment information and materials to potential participants.

Given that this is a quasi-experimental design, the following topics are not applicable: Assignment of interventions: allocation; Sequence generation {16a}; Concealment mechanisms {16b}; Implementation {16c}; Blinding; Who will be blinded {17a}; Procedure for unblinding if needed {17b}.

## DATA COLLECTION AND MANAGEMENT

### Plans for assessment and collection of outcomes {18a}

The study staff member will acquire demographic information and general health history and screen participants for cognitive ability with the Mini-Mental State Examination-short version[64], and for kinesthetic imagery with the Kinesthetic and Visual Imagery questionnaire.[65] All data in this study will be entered into the REDCap database.

### Plans to promote participant retention and complete follow-up {18b}

Participants will be encouraged to complete all scheduled virtual visits and assessments. Given that this study is starting during the COVID-19 pandemic, we estimate 90% adherence and 30% attrition given a high likelihood of some participants being exposed to COVID-19 during the study. If an assessment or weekly check-in is missed, care will be taken to get the information at a later time. There are no stop criteria for missed sessions or testing. The study team will decide when to consider a participant “lost to follow-up” based on clinical/scientific judgment on a case-by-case basis. Missed testing sessions will be recorded for consideration during the analysis but will not be cause for removal from the study.

### Data management {19}

Clinical and weekly check-in data will be entered into the REDCap database, which uses a MySQL database via a secure web interface. During the project, data will be accessible to collaborators through the secure REDCap system by username and password; users are given only the permissions they need to carry out their responsibilities. The MySQL database and the web server will both be housed on secure servers operated by the Academic Health Center’s Information Systems group (AHC-IS). Servers are in a physically secure location on campus and are backed up nightly, with the backups stored in accordance with the AHC-IS retention schedule of daily, weekly, and monthly tapes retained for 1 month, 3 months, and 6 months, respectively. Weekly backup tapes are stored offsite. The AHC-IS servers provide a stable, secure, well-maintained, and high-capacity data storage environment, and REDCap is a widely used, powerful, reliable, well-supported system.

The clinical data on REDCap will be verified and maintained by the study staff and the biostatistician. Source data will be verified for accuracy, and completeness by comparing the data to external data sources (for example, medical records). Data collection forms will all be numerically coded to ensure privacy and data integrity.

### Confidentiality {27}

Research records will be stored in a confidential manner to protect the confidentiality of information of the participants. Participant confidentiality will be safeguarded using password protected databases and locked file cabinets. Research records use the participant code only, with keys identifying individual participants available only to the PI or selected and approved designees. Records of the combined HIPAA-eConsent will be securely stored on the REDCap database server. Further, access to identifiable private information from study participants will only be accessible to study related personnel who have met the UMN’s training requirements for the Responsible Conduct of Research, Good Clinical Practice, HIPAA, and data security and have completed all initial and annual study specific training.

#### Plans for collection, laboratory evaluation and storage of biological specimens for genetic or molecular analysis in this trial/future use {33}

Not applicable, since none such data will be collected in this study.

## STATISTICAL METHODS

### Statistical methods {20a}

All quantitative variables will be summarized using descriptive statistics for all subjects as well as each group at each time point. For longitudinal outcomes, across-time changes and correlations will also be estimated for all participants as well as each group that will be used for sample size determination in future studies. All analyses will be done using standard statistical software including SAS, Stata, and R. The data analysis from this quasi-experimental study will provide an estimate of key trial elements to determine whether to proceed to a larger definitive randomized controlled trial.

The qualitative findings will also serve to reach conclusions on feasibility and acceptability of participating in the study, as well as feasibility, acceptability, and satisfaction of practicing Qigong for adults with SCI-related neuropathic pain.

### Interim analyses {21b}

Stopping rules defined above will be applied. We do not plan to perform an interim analysis.

### Methods for additional analyses (e.g., subgroup analyses) {20b}

Given the limited sample size, we will not consider subgroup analyses at this stage.

### Methods in analysis to handle protocol non-adherence and any statistical methods to handle missing data {20c}

Descriptive statistics and percentages will be calculated for nominal and continuous demographic data and feasibility outcomes. The data will be transformed appropriately to ensure at least approximate normal distributions. Missing data will be handled using the multiple imputation and inverse probability weighting approaches.

### Plans to give access to the full protocol, participant level-data and statistical code {31c}

Data will be stored in the Dryad repository of the UMN and be made available upon request.

## OVERSIGHT AND MONITORING

### Composition of the coordinating center and trial steering committee {5d}

The principal investigator (Dr. Van de Winckel) will be responsible for the fiscal and research administration. Dr. Van de Winckel will oversee study design, study coordination, data management, regulatory compliance, data collection, and analysis. Dr. Morse will oversee recruitment and dissemination activities. The research team will meet over Zoom on a weekly basis to monitor project progress and to trouble-shoot. Publication authorship will be based on the relative scientific contributions of the principal investigator and key personnel.

### Composition of the data monitoring committee, its role and reporting structure **{21a}**

Because there is minimal risk associated with this study, we will not establish a data and safety monitoring board.

### Adverse event reporting and harms {22}

The principal investigator will be responsible for determining whether an adverse event (AE) is expected or unexpected. An AE will be considered unexpected if the nature, severity, or frequency of the event is not consistent with the risk information previously described for the study intervention. We will perform weekly 1-1 automated check-ins throughout the study period when the Qigong intervention is being organized. Participants will also have at least 2 contact numbers and/or e-mail addresses to contact the study team (study staff and principal investigator). Participants will also be instructed to contact the investigators and/or study staff if they experience an adverse event. In case of a serious adverse event the participant is requested to contact the principal investigator immediately. If an event occurs, IRB protocols for adverse event classification and reporting will be followed. In the case of a serious adverse event we will follow the procedure outlined in our IRB plan.

### Frequency and plans for auditing trial conduct {23}

The data integrity monitor from the CTSI at the UMN will visit the site and perform monitoring and data quality assurance biannually, according to the Good Clincal Practice guidelines. The monitor will be responsible for protecting participants from avoidable harm and providing advice to both the funding agency and investigators concerning the scientific and ethical conduct of the trial. The monitor will review study data concerning subject safety, study conduct, and progress.

### Plans for communicating important protocol amendments to relevant parties (e.g. trial participants, ethical committees) {25}

All protocol amendments will be submitted for IRB approval prior to implementation and communication with the participants. If protocol amendments impact the content of the consent form, participants will be reconsented with the new version of the consent form.

### Dissemination plans {31a}

The results will be presented at conferences and in peer-reviewed publications, in which we will acknowledge the stakeholders’ contributions. We will use our existing community connections at the UMN to disseminate information, spread awareness of Qigong and the results, and hold dialogues with stakeholders. This study will help us design future larger studies, strengthen our funding proposals, and provide us with a platform to educate consumers, caregivers, and providers locally and nationally.

## Discussion

In this quasi-experimental study, we will test our hypothesis that Spring Forest Qigong^TM^’s Five Element Qigong Healing Movements is feasible and will reduce SCI-related neuropathic pain by improving body awareness. To our knowledge, this will be the first study using Qigong for neuropathic pain relief in adults with SCI. Adults with SCI-related neuropathic pain were involved in the design of this study. This study will provide preliminary evidence-based data on the feasibility of remotely delivered Qigong and whether Qigong practice reduces neuropathic pain and pain-related outcomes. The remote delivery of Qigong offers multiple applications for broad use in the home or community, as a stand-alone treatment or in conjunction with other treatments.

There are some limitations to this study. This quasi-experimental study will be conducted on a limited sample size and thus the results will need to be validated in a larger sample size and compared to a control group. The follow-up period with no intervention is 6 weeks. We will collect a 1-year follow-up to evaluate possible behavioral change. The data from the present work will inform the design of future randomized controlled trials with adequate sample size, intervention dosage, and follow-up duration.

### Trial status

At the time of protocol submission, we report protocol version 7 and do not anticipate any further changes until trial completion. Recruitment started from July 1, 2021 and the study will run until approximately July, 2023.

## Data Availability

This is a study protocol paper. Therefore it does not contain study data.

## Abbreviations

CTSI: Clinical and Translational Science Institute
IRB: Institutional Review Board
REDCap: Research Electronic Data Capture
SCI: spinal cord injury
UMN: University of Minnesota

## DECLARATIONS

## Acknowledgments

We thank Qigong Grand Master Chunyi Lin, MS, and Qigong Master Jaci Gran from the Spring Forest Qigong^TM^ for their consultancy and assistance with providing the content of the 5 Element Qigong Healing Movements protocol. We thank the adults with SCI and neuropathic pain who gave their valuable input on the research questions, intervention, assessments, and elements of the study design. We present our profound thanks to Marc Noël for the critical review of the manuscript.

## Authors’ contributions {31b}

AVDW, LM, and RB were responsible for the conception and design of the work in collaboration with adults with SCI-related neuropathic pain. AVDW is the principal investigator of the study, oversees, and manages the study. AVDW also communicates with the UMN CTSI’s monitor every 6 months and submits protocol modifications to relevant parties. LM is overseeing participant recruitment. LZ is responsible for statistical analysis. AVDW, SC, and WD will collect the data.

AVDW wrote the original draft. All authors contributed substantially to parts of the manuscript, critically revised it for content, approved the final version, and agreed to be accountable for the accuracy and integrity of this work.

## Funding {4}

This study is internally funded by the Division of Physical Therapy, Department of Rehabilitation Medicine, Medical School, University of Minnesota. The research is supported by the National Institutes of Health’s National Center for Advancing Translational Sciences, grant UL1TR002494. The content is solely the responsibility of the authors and does not necessarily represent the official views of the National Institutes of Health’s National Center for Advancing Translational Sciences. The funders have no role in study design, data collection, and analysis, decision to publish, or preparation of the manuscript.

## Availability of data and materials {29}

The study team will have access to the data set until completion of the study. Upon publication of the data, data will be stored in the Dryad repository of the University of Minnesota and shared upon request.

## Ethics approval and consent to participate {24}

We will adhere to the ethical guidelines of the Declaration of Helsinki. The University of Minnesota’s Institutional Review Board (IRB) approved the study (IRB#STUDY00011997). The study is registered at ClinicalTrial.gov: NCT04917107.

## Consent for publication {32}

We are willing to provide a model eConsent form on request.

## Competing interests {28}

The authors declare that they have no competing interests.

## Notes

### Competing Interest Statement

The authors have declared no competing interest.

### Clinical Trial

NCT04917107

### Author Declarations

The University of Minnesota Institutional Review Board (IRB) approved the study (IRB #STUDY00011997).

### Summary of Updates

We have updated the prevalence data to the most recent evidence: 69% of the 299,000 adults with SCI have neuropathic pain.

